# The Evaluation of Sleep Disturbances for Chinese Frontline Medical Workers under the Outbreak of COVID-19

**DOI:** 10.1101/2020.03.06.20031278

**Authors:** Jing Qi, Jing Xu, Bozhi Li, Jinsha Huang, Yuan Yang, Zhentao Zhang, Dongai Yao, Qunhui Liu, Min Jia, Daokai Gong, Xiaohong Ni, Qimei Zhang, Furong Shang, Nian Xiong, Chunli Zhu, Tao Wang, Xi Zhang

**Author notes:** Corresponding authors: Xi Zhang, Department of Neurology, The Secondary Medical Center, National Clinical Research Center for Geriatric Disease, Chinese PLA General Hospital, Beijing, 100853, China, School of Medicine, Nankai University, Tianjin, 300071, China, Tao Wang, Department of Neurology, Union Hospital, Tongji Medical College, Huazhong University of Science & Technology, Wuhan, Hubei, 430022, China.

## Abstract

**Background:** The outbreak of coronavirus disease 2019 in China remains to be a serious challenge for frontline medical workers (fMW). They are under high risk of being infected and high mental stress, which may lead to sleep disturbances, anxiety, and depression.

**Methods:** We conducted a cross-sectional study to evaluate sleep disturbances of fMW and made a comparison with non-fMW. The medical workers from multiple hospitals in Hubei Province, China, were volunteered to participate. An online questionnaire, including Pittsburgh Sleep Quality Index (PSQI), Athens Insomnia Scale (AIS), and Visual Analogue Scale (VAS), was used to evaluate sleep disturbances and mental status of fMW. Sleep disturbances were defined as PSQI>7 points or/and AIS>6 points. We compared the scores of PSQI, AIS, anxiety and depression VAS, and prevalence of sleep disturbances between fMW and non-fMW. Subgroup analysis for different gender in fMW was conducted.

**Findings:** A total of 1306 subjects (including 801 fMW and 505 non-fMW) were enrolled. Compared to non-fMW, fMW had significantly higher scores of PSQI (p< 0.0001), AIS (p<0.0001), anxiety (p<0.0001) and depression (p=0.0010), and higher prevalence of sleep disturbances with PSQI > 7 points (p<0.0001) and AIS > 6 points (p<0.0001). In subgroup analysis, compared to male fMW, female fMW had significantly higher scores of PSQI (p=0.022) and higher prevalence of sleep disturbances with PSQI > 7 points (p<0.0001).

**Interpretation:** fMW had higher prevalence of sleep disturbances and worse sleep quality than non-fMW. Female fMW were more vulnerable to having sleep disturbances than male fMW.

**Funding:** None.

## Introduction

As a group of enveloped RNA viruses, coronaviruses are distributed broadly among humans and other animals, and can cause multi-organ infections, mainly invade the respiratory system.^1^ In December 2019, several pneumonia patients with cryptogenic etiology were reported in Wuhan, Hubei Province, China.^2^ A novel coronavirus was identified as the arch-criminal and has established efficient human-to-human transmission. It subsequently was named as coronavirus disease 2019 (COVID-19) by the World Health Organization (WHO). With the unknown mighty infectivity, COVID-19 has spread rapidly, confirmed cases were reported in Wuhan, Hubei Province, other regions in China, and other countries successively.^3,4^ The grim situation has attracted worldwide attention. More than 75000 cases were confirmed with COVID-19 in China, including 3000 doctors and nurses at least,^5^ who have become the high-risk susceptible population to the disease. In addition to the risk of being infected with COVID-19, frontline medical workers (fMW) may have sleep disturbances, anxiety, and depression when facing with a magnitude outburst public health incident.

Sleep disturbances are defined as the mental or/and physical status that trigger a series of adverse symptoms owing to abnormal amount of sleep or/and poor sleep quality, and remain to be one of the global health concerns. Sleep disturbances have a prevalence ranging from 8.3% to 45%.^6,7^ By comparison to the general population, it was reported that medical workers may have higher prevalence of insomnia.^8^ Medical workers are generally under great pressure, irregular work time, and frequent day-night work shifts, which may lead to the increasing of sleep disturbances.^9^ In a meta-analysis from China, Qiu et al.^7^ found that 39.2% of Chinese medical workers suffered from sleep disturbances, and the prevalence was much higher than the general population. The outbreak of COVID-19 in China remains to be a serious challenge for fMW. There is no doubt that they are not only under high risk of being infected with the disease but also high mental stress, which may lead to acute sleep disturbances or mental disorders. However, as far as we know, few studies have concentrated on the sleep disturbances of fMW under the emergent events of public health, especially in the outbreak of COVID-19.

Therefore, we conducted a cross-sectional study to evaluate the sleep disturbances of fMW in the battle against COVID-19, and made a comparison with non-fMW.

## Methods

### Study samples

This cross-sectional study included medical workers from multiple hospitals in Hubei Province, China. The Institutional Review Board of Chinese PLA General Hospital approved this study. The study protocol was conducted in accordance with the Declaration of Helsinki. Informed consent was waived due to the cross-sectional nature of this study. All of subjects included in the study were anonymous and volunteered to participate. Subjects were divided into two groups according to the existence of directive exposure to COVID-19 patients, including fMW and non-fMW. The inclusion criteria for this study were as follows: (a) age older than 18 years old; (b) medical workers from hospitals in Hubei Province, including local medical workers and supportive medical workers from other Chinese regions; (c) involved in the battle against COVID-19 directly or/and indirectly, and (d) volunteered to participate in the survey. The exclusion criteria were listed as follows: (a) has been diagnosed as sleep disturbances; (b) sleep quality was affected by physical or psychological diseases; (c) sleep quality was affected by negative personal events; (d) within the treatment course for mental or/and physical diseases, and (e) incomplete data of the online questionnaires.

### Data collection

The online survey was conducted in February 2020 among medical workers from multiple hospitals in Hubei Province, with the contents encompassing basic information (age, gender, marriage, education level, etc.), epidemiological investigation, details of work, Pittsburgh Sleep Quality Index (PSQI), Athens Insomnia Scale (AIS), Visual Analogue Scale (VAS), the factors influencing sleep quality, etc. PSQI is a self-reported questionnaire which assesses subjective sleep quality within one month. It comprises of seven components, including sleep quality, sleep latency, sleep duration, sleep efficiency, sleep disturbances, sleep medication, and daytime function. A subject with total score > 7 points is defined as poor sleep quality.^10^ In general, the higher the PSQI, the worse the sleep quality. AIS is a brief instrument to assess the severity of insomnia. It contains eight items with each item rated from 0 to 3 points. The sum of eight items’ scores is regarded as total score. A subject with total score > 6 points is diagnosed as insomnia. In general, the higher the AIS, the severer the insomnia.^11^ A 10-items VAS measure was used to investigate emotional status of subjects, including work pressure, attention, anxiety, energy, confidence, irritability, stress, impatience, fear, and depression. The VAS was conducted online as a horizontal graphic slider, which was divided into ten segments from one (weakest) to ten (strongest). The VAS scores could represent the degree of their feelings. Higher scores indicate stronger feeling of each emotional state. We used scores of VAS to compare anxiety and depression between fMW and non-fMW.

### Statistical analysis

Categorical variables were described as frequencies and percentages, and continuous variables were described as the mean ± SDs. Statistical analyses were performed using SPSS 23.0 for Windows (IBM, Somers, NY). The demographic characteristics were compared by non-parametric test for continuous variables and by Pearson’s chi-square test for categorical variables between two groups. The scores of PSQI, AIS, anxiety, and depression VAS were compared by ANOVA. The variables with significant differences between two groups in demographic characteristics were described as covariates and joined in the process of ANOVA. In the subgroup analysis of fMW, the scores of PSQI, AIS, anxiety, and depression VAS were compared by ANOVA between male and female fMW. Partial correlation analysis was conducted to investigate the correlation between PSQI, AIS, anxiety, and depression. P value <0.05 was considered to indicate statistical significance in all statistical analyses.

## Results

### Subject demographic characteristics

A total of 1306 medical workers (801 in fMW group and 505 in non-fMW group; Figure 1) with age of (33.1±8.4) years old were enrolled in the study, comprising 256 (19.6%) males and 1050 (80.4%) females. There were 161 (20.1%) males and 640 (79.9%) females in fMW group. The detailed demographic characteristics are presented in Table 1. There were 893 (68.4%) subjects married and 156 (11.9%) subjects had high education level (Master’s degree and Doctor’s degree). Only 148 (11.3%) subjects had advanced-rank in hospitals, who may present with more affluent working experience and positive attitudes toward COVID-19. In demographic characteristics, significant differences of age ([32.4±7.7] vs [34.1±9.3]; p=0.016), education level (p=0.0020), and rank (p=0.0010) were found between the fMW group and non-fMW group.

**Table 1.**
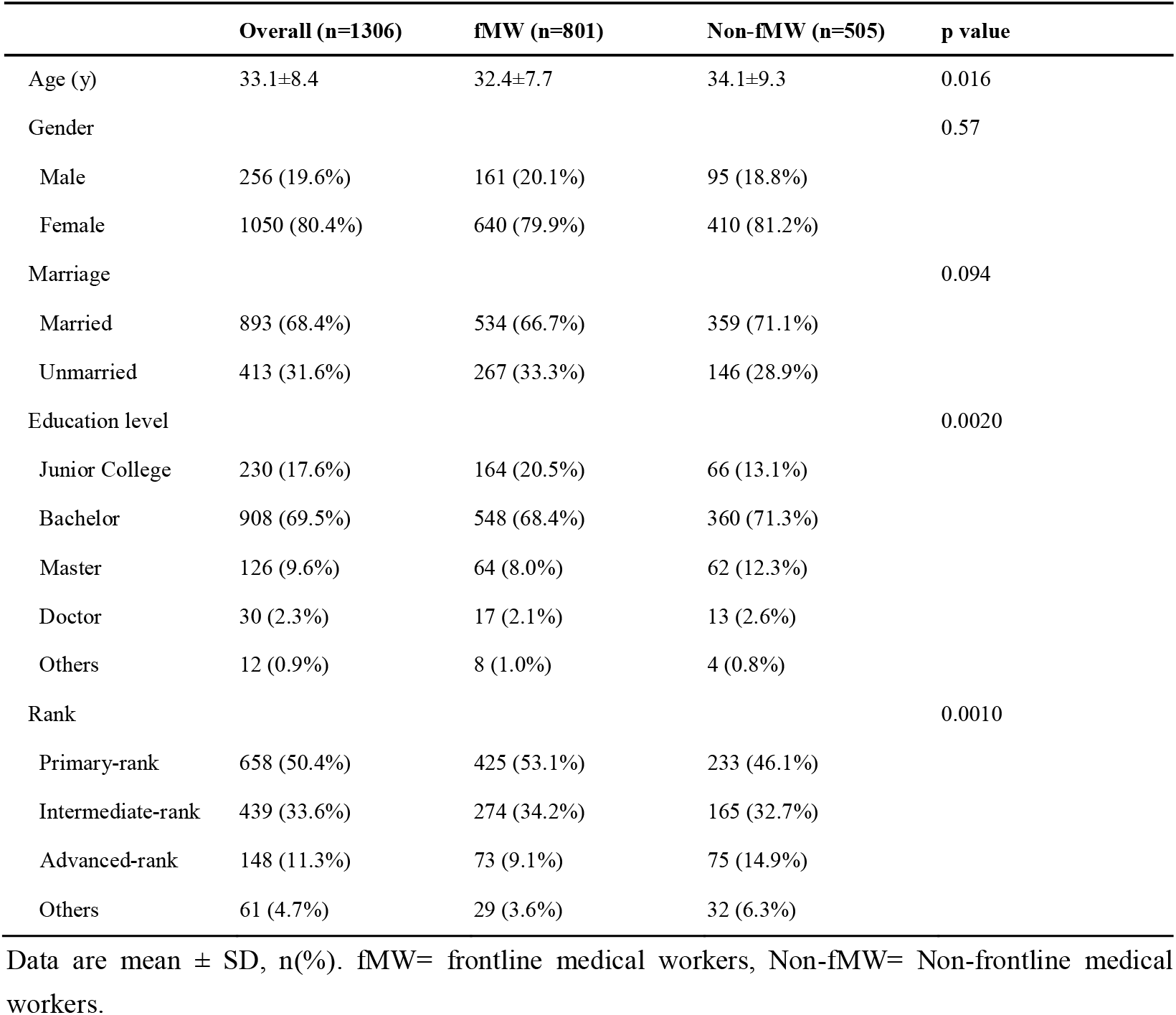
Demographic characteristics of subjects.

|  | Overall (n=1306) | fMW (n=801) | Non-fMW (n=505) | p value |
| --- | --- | --- | --- | --- |
| Age (y) | 33.1±8.4 | 32.4±7.7 | 34.1±9.3 | 0.016 |
| Gender |  |  |  | 0.57 |
| Male | 256 (19.6%) | 161 (20.1%) | 95 (18.8%) |  |
| Female | 1050 (80.4%) | 640 (79.9%) | 410 (81.2%) |  |
| Marriage |  |  |  | 0.094 |
| Married | 893 (68.4%) | 534 (66.7%) | 359 (71.1%) |  |
| Unmarried | 413 (31.6%) | 267 (33.3%) | 146 (28.9%) |  |
| Education level |  |  |  | 0.0020 |
| Junior College | 230 (17.6%) | 164 (20.5%) | 66 (13.1%) |  |
| Bachelor | 908 (69.5%) | 548 (68.4%) | 360 (71.3%) |  |
| Master | 126 (9.6%) | 64 (8.0%) | 62 (12.3%) |  |
| Doctor | 30 (2.3%) | 17 (2.1%) | 13 (2.6%) |  |
| Others | 12 (0.9%) | 8 (1.0%) | 4 (0.8%) |  |
| Rank |  |  |  | 0.0010 |
| Primary-rank | 658 (50.4%) | 425 (53.1%) | 233 (46.1%) |  |
| Intermediate-rank | 439 (33.6%) | 274 (34.2%) | 165 (32.7%) |  |
| Advanced-rank | 148 (11.3%) | 73 (9.1%) | 75 (14.9%) |  |
| Others | 61 (4.7%) | 29 (3.6%) | 32 (6.3%) |  |

Data are mean ± SD, n(%). fMW= frontline medical workers, Non-fMW= Non-frontline medical workers.

**Figure 1.**
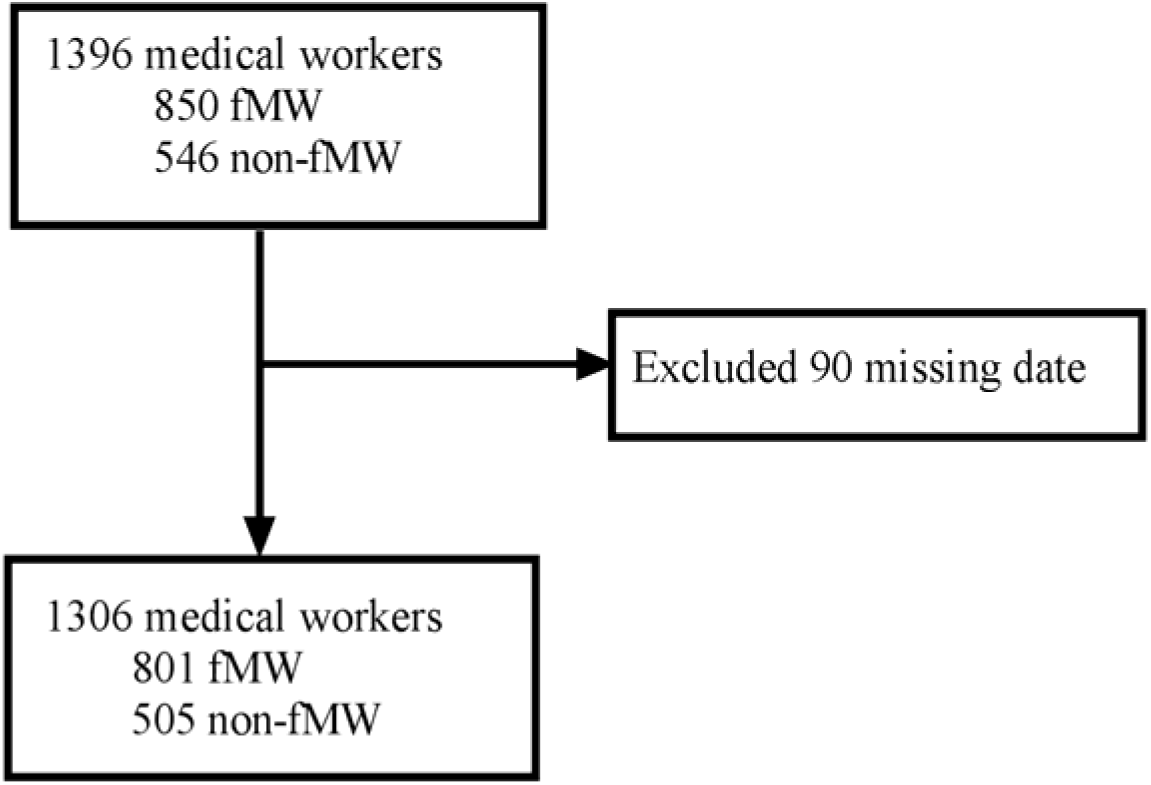
The flowchart shows the subject selection. fMW= frontline medical workers, non-fMW= non-frontline medical workers.

### Comparison of scores and prevalence

The detailed comparisons of scores between two groups are presented in Table 2. The scores of PSQI, AIS, anxiety, and depression VAS in fMW group were 9.3±3.8, 6.9±4.3, 4.9±2.7, 4.1±2.5, respectively, and in non-fMW group, they were 7.5±3.7, 5.3±3.8, 4.3±2.6, 3.6±2.4, respectively. Compared to non-fMW group, the fMW group had significantly higher scores of PSQI (9.3 vs 7.5; p<0.0001; Figure 2A), AIS (6.9 vs 5.3; p<0.0001; Figure 2B), anxiety (4.9 vs 4.3; p<0.0001; Figure 2C), and depression (4.1 vs 3.6; p=0.0010; Figure 2D).

**Table 2.** Comparisons of PSQI, AIS, Anxiety and Depression VAS scores between fMW and non-fMW.

|  | Overall (n=1306) | fMW (n=801) | Non-fMW (n=505) | p value |
| --- | --- | --- | --- | --- |
| PSQI | 8.6±3.9 | 9.3±3.8 | 7.5±3.7 | < 0.0001 |
| AIS | 6.3±4.2 | 6.9±4.3 | 5.3±3.8 | < 0.0001 |
| Anxiety VAS | 4.7±2.7 | 4.9±2.7 | 4.3±2.6 | < 0.0001 |
| Depression VAS | 3.9±2.4 | 4.1±2.5 | 3.6±2.4 | < 0.0001 |

Data are mean ± SD. fMW= frontline medical workers, Non-fMW= Non-frontline medical workers, PSQI= Pittsburgh Sleep Quality Index, AIS= Athens Insomnia Scale, VAS= Visual Analogue Scale.

**Figure 2.**
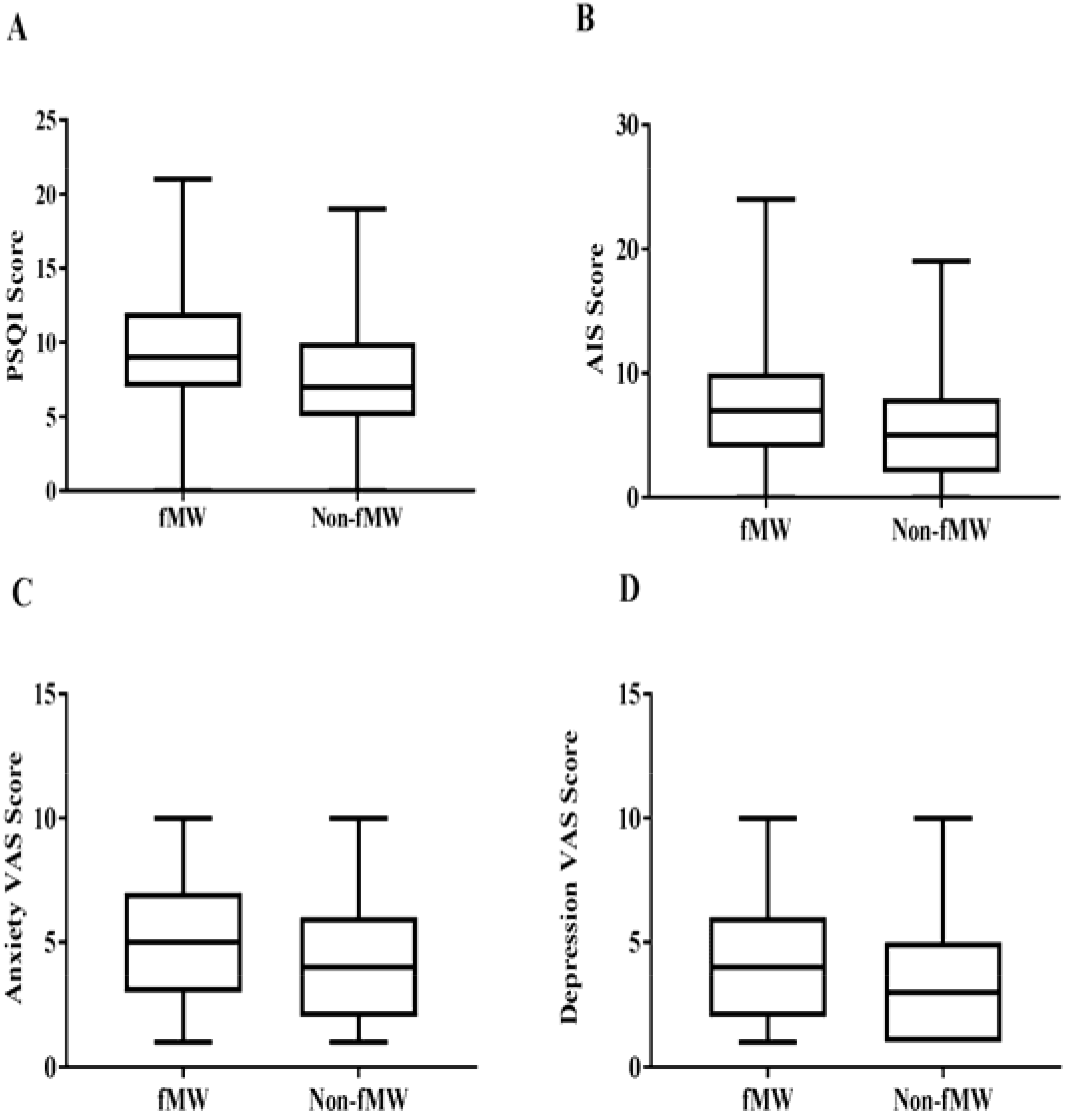
The boxplots for PSQI, AIS, Anxiety and Depression VAS scores of fMW and non-fMW. (A) The fMW had significantly higher scores of PSQI (9.3 vs 7.5; p< 0.0001) than non-fMW. (B) The fMW had significantly higher scores of AIS (6.9 vs 5.3; p< 0.0001) than non-fMW. (C) The fMW had significantly higher scores of anxiety VAS (4.9 vs 4.3; p< 0.0001) than non-fMW. (D) The fMW had significantly higher scores of depression VAS (4.1 vs 3.6; p=0.0010) than non-fMW. PSQI= Pittsburgh Sleep Quality Index, AIS= Athens Insomnia Scale, VAS= Visual Analogue Scale, fMW= frontline medical workers, Non-fMW= Non-frontline medical workers.

The detailed comparison of prevalence of sleep disturbances between two groups are presented in Table 3. There were 538 (67.2%) of 801 fMW and 241 (47.7%) of 505 non-fMW with PSQI > 7 points, which indicated over 60% of fMW had poor sleep quality. Meanwhile, AIS > 6 points was observed in 414 (51.7%) of 801 fMW and 180 (35.6%) of 505 non-fMW, which indicated that over half of fMW had insomnia. Furthermore, compared to non-fMW group, the fMW group had significantly higher prevalence of sleep disturbances, according to PSQI > 7 points (538 [67.2%] of 801 vs 241 [47.7%] of 505; p<0.0001; Figure 3) and AIS > 6 points (414 [51.7%] of 801 vs 180 [35.6%] of 505; p<0.0001; Figure 3).

**Table 3.** Comparison of prevalence of sleep disturbances between fMW and non-fMW.

|  | Overall (n=1306) | fMW (n=801) | Non-fMW (n=505) | p value |
| --- | --- | --- | --- | --- |
| PSQI>7 | 779 (59.6%) | 538 (67.2%) | 241 (47.7%) | <0.0001 |
| AIS>6 | 594 (45.5%) | 414 (51.7%) | 180 (35.6%) | <0.0001 |

Data are n(%). fMW= frontline medical workers, Non-fMW= Non-frontline medical workers, PSQI= Pittsburgh Sleep Quality Index, AIS= Athens Insomnia Scale.

**Figure 3.**
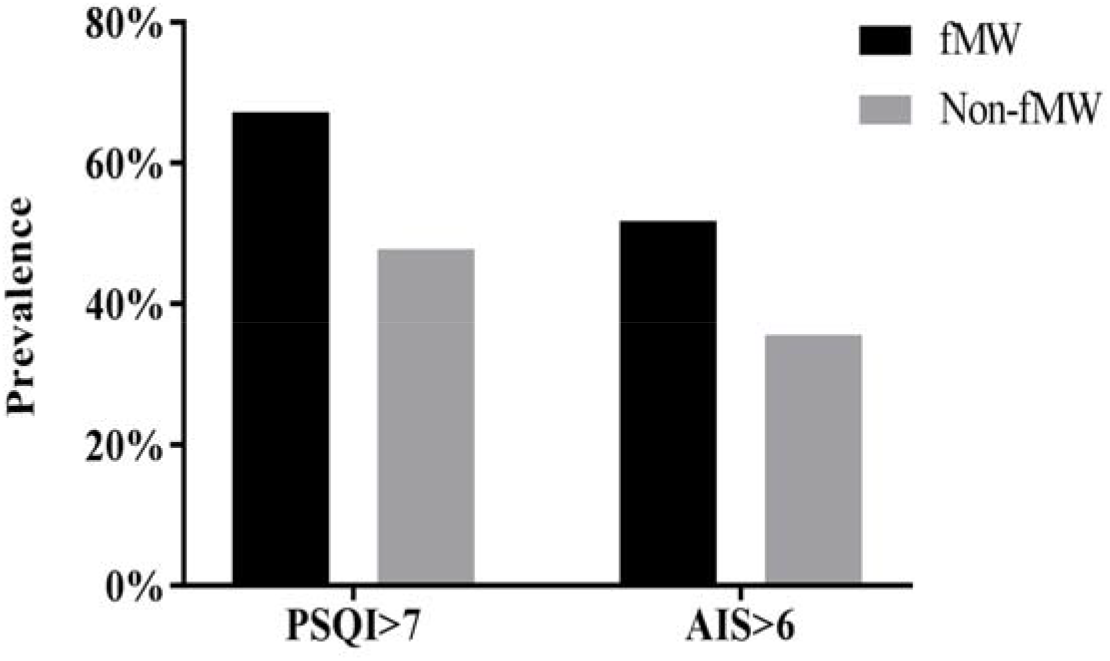
The prevalence of sleep disturbances of fMW and non-fMW. The fMW group had significantly higher prevalence of sleep disturbances than non-fMW, according to PSQI > 7 points (538 [67.2%] of 801 vs 241 [47.7%] of 505; p<0.0001) and AIS > 6 points (414 [51.7%] of 801 vs 180 [35.6%] of 505; p<0.0001). PSQI= Pittsburgh Sleep Quality Index, AIS= Athens Insomnia Scale, fMW= frontline medical workers, Non-fMW= Non-frontline medical workers.

In the subgroups analysis, detailed comparisons of scores between male and famale fMW are presented in Table 4. The scores of PSQI, AIS, anxiety, and depression VAS in male fMW group were 8.6±4.3, 6.4±4.7, 4.6±2.7, 3.8±2.4, respectively, and in female fMW group, they were 9.4±3.8, 7.0±4.2, 5.0±2.7, 4.2±2.5, respectively. Compared to the male fMW group, female fMW group had significantly higher score of PSQI (9.4 vs 8.6; p=0.022; Figure 4A), while no significant differences were found in AIS (7.0 vs 6.4; p=0.11), anxiety (5.0 vs 4.6; p=0.14), and depression (4.2 vs 3.8; p=0.20). In addition, 450 (70.3%) of 640 female fMW and 88 (54.7%) of 161 male fMW had PSQI >7 points. And compared to male fMW group, the female fMW group had significantly higher prevalence of sleep disturbances, according to PSQI > 7 points (450 [70.3%] of 640 vs 88 [54.7%] of 161; p*<*0.0001; Figure 4B). In summary, the female fMW in battle against COVID-19 are more vulnerable to suffering from sleep disturbances according to the PSQI score.

**Table 4.** Comparisons of PSQI, AIS, Anxiety and Depression VAS scores between male and female fMW.

|  | Overall (n=801) | Male fMW (n=161) | Female fMW (n=640) | p value |
| --- | --- | --- | --- | --- |
| PSQI | 9.3±3.8 | 8.6±4.3 | 9.4±3.8 | 0.022 |
| AIS | 6.9±4.3 | 6.4±4.7 | 7.0±4.2 | 0.11 |
| Anxiety VAS | 4.9±2.7 | 4.6±2.7 | 5.0±2.7 | 0.14 |
| Depression VAS | 4.1±2.5 | 3.8±2.4 | 4.2±2.5 | 0.20 |

Data are mean ± SD. fMW= frontline medical workers, PSQI= Pittsburgh Sleep Quality Index, AIS= Athens Insomnia Scale, VAS= Visual Analogue Scale.

**Figure 4.**
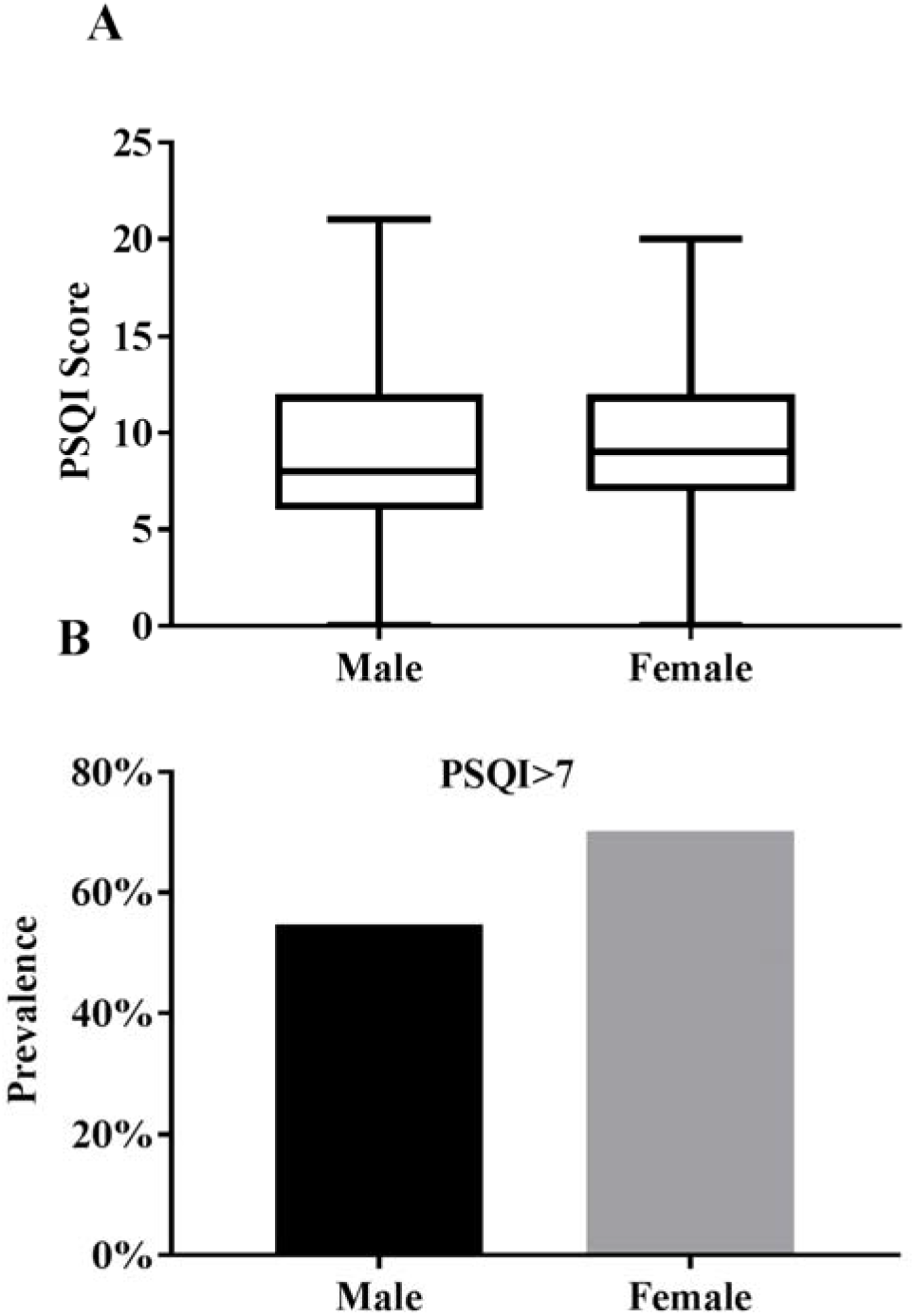
The score of PSQI and prevalence of sleep disturbances of male fMW and female fMW. (A) The female fMW had significantly higher scores of PSQI (9.4 vs 8.6; p=0.022) than male fMW. (B) The prevalence of sleep disturbances was significantly higher in female fMW than male fMW (450 [70.3%] of 640 vs 88 [54.7%] of 161; p*<*0.0001) according to PSQI>7. PSQI= Pittsburgh Sleep Quality Index.

### The factors influencing sleep quality

The detailed comparisons of factors influencing sleep quality between fMW group and non-fMW group are presented in Table 5. From the perspective of 1306 medical workers, severity of COVID-19 was the leading factor (497 [38.1%]) influencing sleep quality, followed by frequent work shifts (415 [31.8%]), work stress (408 [31.2%]), and insufficient sleep time (235 [18.0%]). Compared to non-fMW, more fMW believed work stress (305 [38.1%] of 801 vs 103 [20.4%] of 505; p<0.0001; Figure 5), frequent work shifts (320 [40.0%] of 801vs 95 [18.8%] of 505; p<0.0001; Figure 5), and severity of COVID-19 (338 [42.2%] of 801 vs 159 [31.5%] of 505; p< 0.0001; Figure 5) may influence their sleep quality.

**Table 5.**
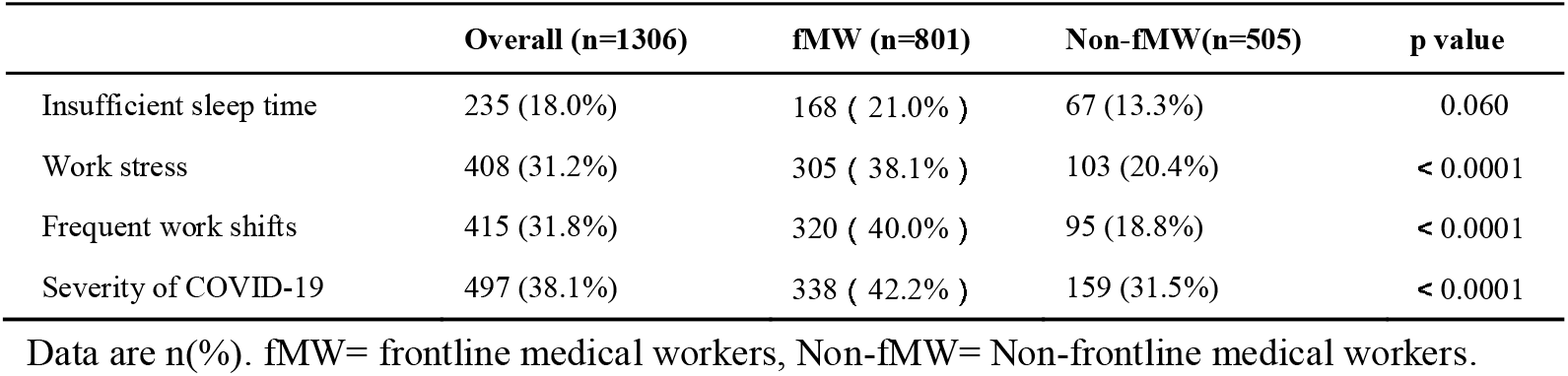
The factors influencing sleep quality from the perspective of medical workers.

|  | Overall (n=1306) | fMW (n=801) | Non-fMW(n=505) | p value |
| --- | --- | --- | --- | --- |
| Insufficient sleep time | 235 (18.0%) | 168 ( 21.0% ) | 67 (13.3%) | 0.060 |
| Work stress | 408 (31.2%) | 305 ( 38.1% ) | 103 (20.4%) | < 0.0001 |
| Frequent work shifts | 415 (31.8%) | 320 ( 40.0% ) | 95 (18.8%) | < 0.0001 |
| Severity of COVID-19 | 497 (38.1%) | 338 ( 42.2% ) | 159 (31.5%) | < 0.0001 |

Data are n(%). fMW= frontline medical workers, Non-fMW= Non-frontline medical workers.

**Figure 5.**
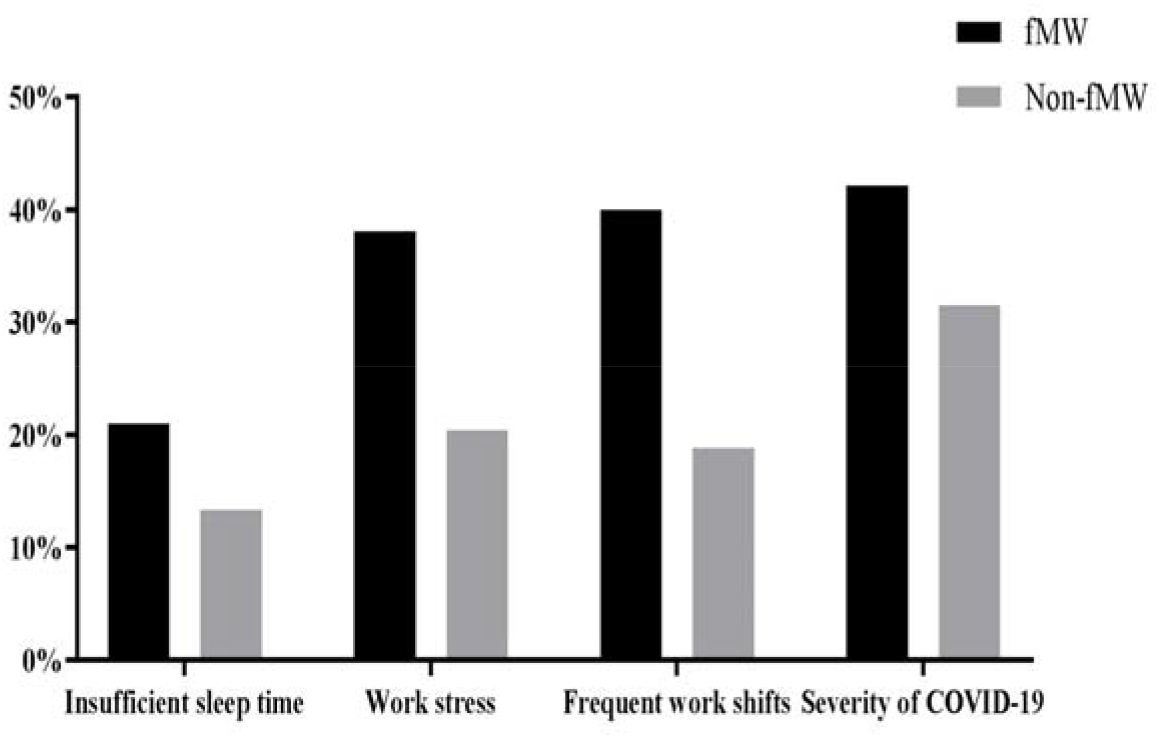
The percentage of factors influencing sleep quality of fMW and non-fMW. fMW= frontline medical workers, Non-fMW= Non-frontline medical workers.

### Partial correlation analysis of PSQI, AIS, anxiety, and depression VAS

Partial correlation analysis was conducted to explore association between PSQI, AIS, anxiety, and depression VAS by adjusting for gender (Table 6). PSQI was significantly correlated with anxiety (r_p_=0.442; p<0.0001) and depression (r_p_=0.329; p<0.0001). And significant association was observed between AIS and anxiety (r_p_=0.526; p<0.0001), and depression (r_p_=0.415; p<0.0001).

**Table 6.** Partial correlation analysis of PSQI, AIS, Anxiety and Depression VAS.

|  | Anxiety VAS |  | Depression VAS |  |
| --- | --- | --- | --- | --- |
| | $r_p$ | p value | $r_p$ | p value |
| PSQI | 0.442 | <0.0001 | 0.329 | <0.0001 |
| AIS | 0.526 | <0.0001 | 0.415 | <0.0001 |

Partial correlation analysis was adjusted for gender. $r_p$ = correlation coefficient, PSQI= Pittsburgh Sleep Quality Index, AIS= Athens Insomnia Scale, VAS= Visual Analogue Scale.

## Discussion

The outbreak of COVID-19 in China has aroused extensive public concern in recent months.^2^ Many countries are also experiencing the invasion of COVID-19, such as Japan, South Korea, Italy, Iran, etc.^12^ The rapid spread of the disease and inadequate early realization toward COVID-19 challenged the health institutions in many countries. In addition, the mortality of COVID-19 was 2.3%, and medical workers accounted for 3.8% in COVID-19 patients, as reported by the latest epidemiological study.^13^ Previous studies have indicated that in ordinary times, medical workers were vulnerable to sleep disturbances, which may be higher than the general population.^7,8^ Therefore, we hypothesized that fMW under the outbreak of COVID-19 in China may be more susceptible to sleep disturbances. To the best of our knowledge, this is the first study to evaluate the sleep disturbances of fMW under the outbreak of COVID-19.

Although sleep disturbances distribute widely among the population, the prevalence varies among different occupations and work pressure.^14-16^ A USA National Health Interview Survey showed short sleep duration disturbed 29.9% of civilian employed workers, including 40.5% of enterprise managers, 37.1% of transportation/warehousing, and 34.8% of manufacturing, and indicated that sleep disturbances have increased over the past decades.^15^ Similarly, a nationwide study in China demonstrated that civil servants had the shortest sleep duration (Mean 7.85 hours) and the worst sleep quality, who may experience with more work pressure.^14^ Meanwhile, 64.1% of police officers had PSQI > 5 points, and were regarded as sleep disturbances as reported. Their sleep disturbances were related to posttraumatic stress and general psychopathology.^17^ A survey from Japan indicated that high occupational stress was significantly associated with insomnia, especially the presence of high efforts but low reward.^18^ And exposure to long working hours and irregular work shifts may attribute to stress, fatigue, and chronic diseases,^19^ which may influence the quantity and quality of sleep. Schiller et al.^16^ compared sleep quality and sleep duration between full worktime subjects and 75% reduced worktime subjects, and found the latter presented with improved sleep quality and sleep duration. The reduction of worktime may be useful for alleviating sleep disturbances.

Accompanied with long working hours, frequent day-night shifts, excessive workload, and high stress, medical workers were vulnerable to sleep disturbances or sleep deprivation all over the world.^20^ Pikovsky et al.^21^ found that 63% and 49% of medical residents presented with low professional performance and judgement levels after the night shifts, respectively. The long working hours could potentially increase the fatigue of medical workers and be linked to the occurrence of medical errors. ^22^ Meanwhile, sleep deprivation could influence the medical workers in making quick and appropriate decisions for patients, which was fatal to patient care.^23^ Deng et al. ^24^ reported that job difficulty, doctor-patient relationship, psychosomatic state, environment or events, promotion or competition, and total pressure scores were related to sleep disturbances for community nurses. Tucker et al.^25^ found moderate work time control for medical workers could improve the sleep quantity that frequent nigh work resulted in, and work time control had association with fewer sleep disturbances. Therefore, it is rational to believe that increased work stress plays a critical role in sleep disturbances of medical workers. With the booming of Chinese economy in past decades, patient visits per physician in China increased by 135% and inpatient admissions per physician rose by 184%. Workload has increased dramatically for Chinese physicians, which exerted a trend that potentially threatened physicians’ health and quality of patient care.^26^ The prevalence of sleep disturbances in Chinese medical workers was 12.9%-78%.^7^ The present study showed that over 50% of fMW under the outbreak of COVID-19 had sleep disturbances. Although it is even higher than that of in normal times, the prevalence is still compatible to previous results from China. The high prevalence of sleep disturbances of fMW in battle against COVID-19 should be put much emphasize. It was reported that sleep characteristics may exert critical effects on immune functions, which may increase the inflammatory response to sleep deprivation and exert negative feedback to aggravate the short sleep duration and increase the risk of cardiovascular, respiratory, and metabolic disorders. ^27^ Thus, sleep disturbances would further increase the possibility of fMW being infected with COVID-19 owing to affected immune system. Therefore, it is imperative to concern on sleep disturbances of fMW, aiming to maintain their healthy condition and guarantee their professional performance in the battle against COVID-19.

Our study showed that the overall scores of PSQI and AIS for fWM were significantly higher than that of non-fWM, which indicated fMW had worse sleep compared to non-fMW. There were 538 (67.2%) and 414 (51.7%) of 801 fMW with PSQI > 7 points and AIS > 6 points, respectively, which were regarded as having sleep disturbances. Our study also demonstrated that the fWM group had significantly higher prevalence of sleep disturbances than that of non-fWM. In addition, the fWM group had significantly longer sleep latency, shorter sleep duration, lower sleep efficiency, and worse daytime function according to the scores of components of PSQI. Change of work environment may underpin the severer sleep disturbances of fMW. With the direct contact with COVID-19 patients, fMW are inclined to emerge anxiety and worries of being infected. The rapid spread of COVID-19 boomed the medical demands, aggravated the shortage of medical resources, and increased work stress for fMW, especially for those with continuous working and frequent day-night shifts. And in terms of factors influencing sleep quality, severity of COVID-19, frequent work shifts, and work stress were the leading factors according to our study. We further found anxiety and depression were significantly correlated with PSQI and AIS in fMV, which indicated that anxious and depressive symptoms may influence sleep quality of fMW. The result was consistent with previous studies demonstrating that anxiety and depression played a key role in the development of sleep disturbances.^28^ This would provide evidence for further targeted interventions to improve sleep disturbances in fMW. However, with the import of supportive fMW from other regions of China, the work duration of fMW may be decreased. By comparison, the non-fMW worked in a relatively mild environment and had less possibility of being infected, thus having less sleep disturbances. Our study also found that scores of PSQI among female fMW were significantly higher than that of male fMW although other scores showed no significant difference, which indicated that female fMW had worse sleep quality than male fMW in emergent events of public health. In a meta-analysis, Zhang et al. ^29^ calculated a hazard ratio of 1.41 for female versus male toward insomnia. In general, females had inferior symptom bearing and more bodily vigilance, and the social culture encouraged females to express the indisposition more,^30^ which potentially exaggerated the severity and prevalence of sleep disturbances. In addition, females tended to report more sleep problems even if having a similar degree of morning awakening compared to males.^29^

Our study has several limitations that should be noted. First, this is a cross-sectional study. All subjects were volunteered to participate in the survey, so there may be subject selection bias. Second, other occupations who directly expose to COVID-19 patients may experience sleep disturbances potentially, such as police and social workers, but our study merely included the medical workers. Third, our questionnaires did not contain sufficient items, such as the potential risk factors for sleep disturbances of fMW, which can provide more information and be used to explore the underlying mechanisms. Forth, our study lacked of interventions and follow-ups, which may show the following changes of sleep disturbances for fMW.

In conclusion, our study showed that over half of the fMW had sleep disturbances in the outbreak of COVID-19. fMW had higher prevalence of sleep disturbances and worse sleep quality than non-fMW. And female fMW showed severer sleep disturbances compared to male fMW. Further interventions should be administrated for fMW, aiming to maintain their healthy condition and guarantee their professional performance in the battle against COVID-19. However, further prospective randomized controlled trials are warranted to validate our results.

## Data Availability

No additional data available.

## Contributors

Jing Qi did the study design, data collection, date interpretation, statistical analysis, and wrote the first draft. Jing Xu and Bozhi Li participated in the study design, did the data collection, date interpretation, statistical analysis, and prepared the figures. Jinsha Huang, Yuan Yang, Zhentao Zhang, Dongai Yao, Qunhui Liu, Min Jia, Daokai Gong, Xiaohong Ni, Qimei Zhang, Furong Shang, Nian Xiong, and Chunli Zhu did the date collection. Xi Zhang and Tao Wang designed the study and revised the manuscript. All authors approved the final version of the manuscript.

